# SARS-CoV-2 mRNA Vaccination-Associated Myocarditis in Children Ages 12-17: A Stratified National Database Analysis

**DOI:** 10.1101/2021.08.30.21262866

**Authors:** Tracy Beth Høeg, Allison Krug, Josh Stevenson, John Mandrola

## Abstract

**Objectives:** Establishing the rate of post-vaccination cardiac myocarditis in the 12-15 and 16-17-year-old population in the context of their COVID-19 hospitalization risk is critical for developing a vaccination recommendation framework that balances harms with benefits for this patient demographic.

**Design, Setting and Participants:** Using the Vaccine Adverse Event Reporting System (VAERS), this retrospective epidemiological assessment reviewed reports filed between January 1, 2021, and June 18, 2021, among adolescents ages 12-17 who received mRNA vaccination against COVID-19. Symptom search criteria included the words chest pain, myocarditis, pericarditis and myopericarditis to identify children with evidence of cardiac injury. The word troponin was a required element in the laboratory findings. Inclusion criteria were aligned with the CDC working case definition for probable myocarditis. Stratified cardiac adverse event (CAE) rates were reported for age, sex and vaccination dose number. A harm-benefit analysis was conducted using existing literature on COVID-19-related hospitalization risks in this demographic.

**Main outcome measures:** 1) Stratified rates of mRNA vaccine-related myocarditis in adolescents age 12-15 and 16-17; and 2) harm-benefit analysis of vaccine-related CAEs in relation to COVID-19 hospitalization risk.

**Results:** A total of 253 CAEs were identified. Rates per million following dose 2 among males were 162.2 (ages 12-15) and 93.0 (ages 16-17); among females, rates were 13.0 and 12.5 per million, respectively. For boys 12-15 without medical comorbidities receiving their second mRNA vaccination dose, the rate of CAE is 2.6 to 4.3 times higher than their 120-day COVID-19 hospitalization risk even at times of peak incidence such as during the delta wave (7-day hospitalizations 2.1/100k population). For boys 16-17 without medical comorbidities, the rate of CAE is 1.5 to 2.5 times higher at times of high weekly COVID-19 hospitalization.

**Conclusions:** Post-vaccination CAE rate was highest in young boys aged 12-15 following dose two. For boys 12-17 without medical comorbidities, the likelihood of post vaccination dose two CAE is 162.2 and 93.0/million respectively. This incidence exceeds their expected 120-day COVID-19 hospitalization rate at both moderate and high COVID-19 hospitalization incidence. Further research into the severity and long-term sequelae of post-vaccination CAE is warranted. Quantification of the benefits of the second vaccination dose and vaccination in addition to natural immunity in this demographic may be indicated to minimize harm.

## INTRODUCTION

Pfizer-BioNTech BNT162b2 and Moderna mRNA-1273 vaccines for SARS-CoV-2 have demonstrated exceptional safety and real-world effectiveness in preventing severe disease and death from COVID-19. Concerns about vaccination-related myocarditis in young men were initially raised in Israel with rates between <u>1/3000-1/6000</u>.[1] In the United States, the initial <u>Centers for Disease Control and Prevention (CDC) report</u>[2,3] identified a rate of approximately 12.6 per million (1/80,000) second doses administered in ages 12-39, but approximately 1/15,000 for males 12-17 and 1/19,000 for males 18-24.[2,3]

Although the CDC analyses [2,3] identified a higher rate of myocarditis in boys than young men, further stratification by adolescent age group (e.g., 12-15 and 16-17 years) was not provided. A second potential limitation was the sensitivity of the CDC symptom search inclusion criteria, which may have failed to identify cases of cardiac adverse events (CAEs), consistent with myocarditis, with objective evidence of cardiac injury following vaccination.

On August 23^rd^, the Federal Drug Administration (FDA) released a Pfizer-BioNTech vaccine report[4] which outlines “an excess risk [of myocarditis] approaching 200 cases/million” or 1/5000 in 16–17-year-old boys, which was three times higher than reported by the CDC [2,3]. In their harm-benefit analysis, the most likely scenario was the benefits of vaccination would outweigh harms in 16-17-year-old males, but “predicted excess cases of vaccine-associated myocarditis/pericarditis would exceed COVID-19 hospitalizations and deaths under the ‘worst case’ scenario.” [4]

Post-vaccination myocarditis rates for the 12-15-year-old age group had not yet been reported beyond the initial trial with 1131 vaccination recipients [5] despite more than 1.2 million children receiving a second dose by mid-June.[6] On August 30^th^, 2021, the CDC reported [7] rates for boys ages 12-15 and 16-17 within seven days of their second dose of Pfizer-BioNTech mRNA vaccination as follows: 42.6 and 71.5 per million, respectively. For females, rates were 4.3 and 8.1 per million, respectively. A harm-benefit analysis of vaccination in the pediatric age group has not yet been performed based on presence of absence of underlying medical conditions. While medical comorbidities confer higher risk for severe COVID-19, this is not known to be the case for post-vaccination myocarditis.

The CDC [2,7] reported a 94-96% hospitalization rate for VAERS-identified myocarditis. The CDC used a 120-day COVID-19 hospitalization rate as a meaningful comparator to vaccination-related harms, and we have chosen to use this same comparison for our study.

Our primary aim was to stratify post-mRNA vaccination myocarditis by age and vaccination dose within the 12–17-year-old population. Our secondary aim was to provide an updated estimate to complement the CDC’s [2,3,7] and FDA’s [4] findings. Our final aim was to perform a harm-benefit analysis of mRNA COVID-19 vaccination myocarditis with that of COVID-19 hospitalization for children with and without one or more comorbidity at low, moderate, and high 120-day COVID-19 hospitalization rates.

## METHODS

We searched the Vaccine Adverse Event Reporting System (VAERS) data for females and males ages 12-17 in reports processed from 1/1/2021 through 6/18/2021 with diagnoses of “myocarditis,” “pericarditis,” “myopericarditis” or “chest pain” in the symptom notes and required the term “troponin” in the laboratory data. We defined a CAE using the CDC working case definition for a probable case.[2] Specifically, the symptom of “chest pain” required at least one of the following: diagnosis of myocarditis, peri- or myopericarditis, acute myocardial infarction; elevated troponin; abnormal electrocardiogram (EKG), abnormal echocardiogram (ECHO), or cardiac MRI (cMRI) findings consistent with myocarditis (as defined in Supplement 1). Cases and hospitalizations with an unknown dose number were assigned to dose 1 or dose 2 in the same proportion as the known doses: 15% occurred following dose 1 and 85% occurred following dose 2.

To compute crude rates per million for doses 1 and 2, our denominators included all children with at least 1 dose of any vaccination and all fully vaccinated children, respectively, as of 6/11/2021[6] to accommodate both reporting lag and a pre-defined 7-day risk window, consistent with the CDC’s analysis. 95% Poisson confidence intervals were calculated for these rates.

To perform a harm-benefit analysis, pediatric hospitalization rates for COVID-19 were obtained from the CDC’s COVID-NET.[8] COVID-19 hospitalization rates among children with and without one or more medical comorbidities were calculated based on hospitalization rates at times of low (June 2021), moderate (August 2021) and high (January 2021) incidence.[8] Children with at least one medical comorbidity were considered to have 4.8 times the likelihood of COVID-19 hospitalization as those without comorbidities based on 70% of children hospitalized for COVID-19 having one or more medical comorbidity.[9] We estimate 33% of children in this age group have one or more comorbidity based upon current data[10] suggesting 21.2% of children 12-19 have obesity and around 8.4% have asthma.[11] These comorbidities are also found in the summary of underlying conditions for pediatric COVID-19 hospitalizations reported to COVID-NET.[8] The two most common underlying conditions among pediatric hospitalizations are obesity (33.8%) and asthma (14.8%).[8] Other relevant comorbidities such as diabetes[11] or the medically complex children[12] appear to make up <5% of this demographic. The estimated ratio of expected hospitalizations for children by presence or absence of comorbidity must therefore account for the relative proportions of children with and without comorbidities admitted as inpatients (70% to 30%, or 2.33:1) compared to their representation in the population (33% to 67%, or 0.49:1). Thus, comparing these two prevalence ratios, 2.33 to 0.49, the odds of hospitalization was considered 4.8-fold higher for children with at least one medical comorbidity.

We provided an additional rate which adjusted for the reported approximate 40% of pediatric hospitalizations for COVID-19 being incidental positive tests on admission.[9,13,14] Finally, the risk of post-vaccination CAE post-dose two in 12–15-year-old-boys was compared with their overall risk of hospitalization from COVID-19 according to presence or absence of comorbidities and adjusted for the 40% hospitalization overestimate as well as the asymptomatic fraction of pediatric cases.

Data were analyzed using Microsoft PowerBI, Stata and Microsoft Excel

## RESULTS

A total of 276 reports met our initial search criteria; of these, 22 were excluded because they had no objective evidence of elevated troponins or abnormal findings on ECG/EKG or ECHO, or we could not exclude the possibility of viral myocarditis or concomitant pneumonia. Of the 22 excluded cases, nine were hospitalized. Ten were 12-15 and twelve were 16-17 years old. Four cases were excluded despite being coded “myocarditis” or “pericarditis” because we could not exclude viral etiology. Of the remaining 254 cases, we excluded from sex-specific rates one case with “unknown” sex, leaving 253 cardiac adverse event reports for full analysis (23 females and 230 males). Of these, 208 cases had peak troponin values available for analysis (Supplement 2); 37 reports did not include the vaccine dose number. Of the 253 included cases, 252 had received Pfizer-BioNTech mRNA vaccination, one had received Moderna mRNA vaccination (only Pfizer-BioNTech was approved for vaccination of children <18 at the time of our database search).

The CAE rates by age and sex and vaccination dose are shown in Figure 1 and Table 1. (Interactive data visualizations and full VAERS case notes available at this link: https://bit.ly/Krug-MyoPerdicarditis). Our post-second-dose-vaccination rates of CAE among adolescent boys aged 12-15 was 162.2/million, 3.8 times higher than the rate reported by the CDC (42.6/million)[7]. Among boys age 16-17, our estimate was 93.0/million, 30% higher than the CDC estimate of 71.5/million. For girls 12-15 years old, our rate was 13.0/million, which was 3 times higher that the CDC’s estimate of 4.3/million.[7] Among girls 16-17, our estimate was 12.5/million, which was 54% higher than the CDC’s estimate of 8.1/million.

**Figure 1.**
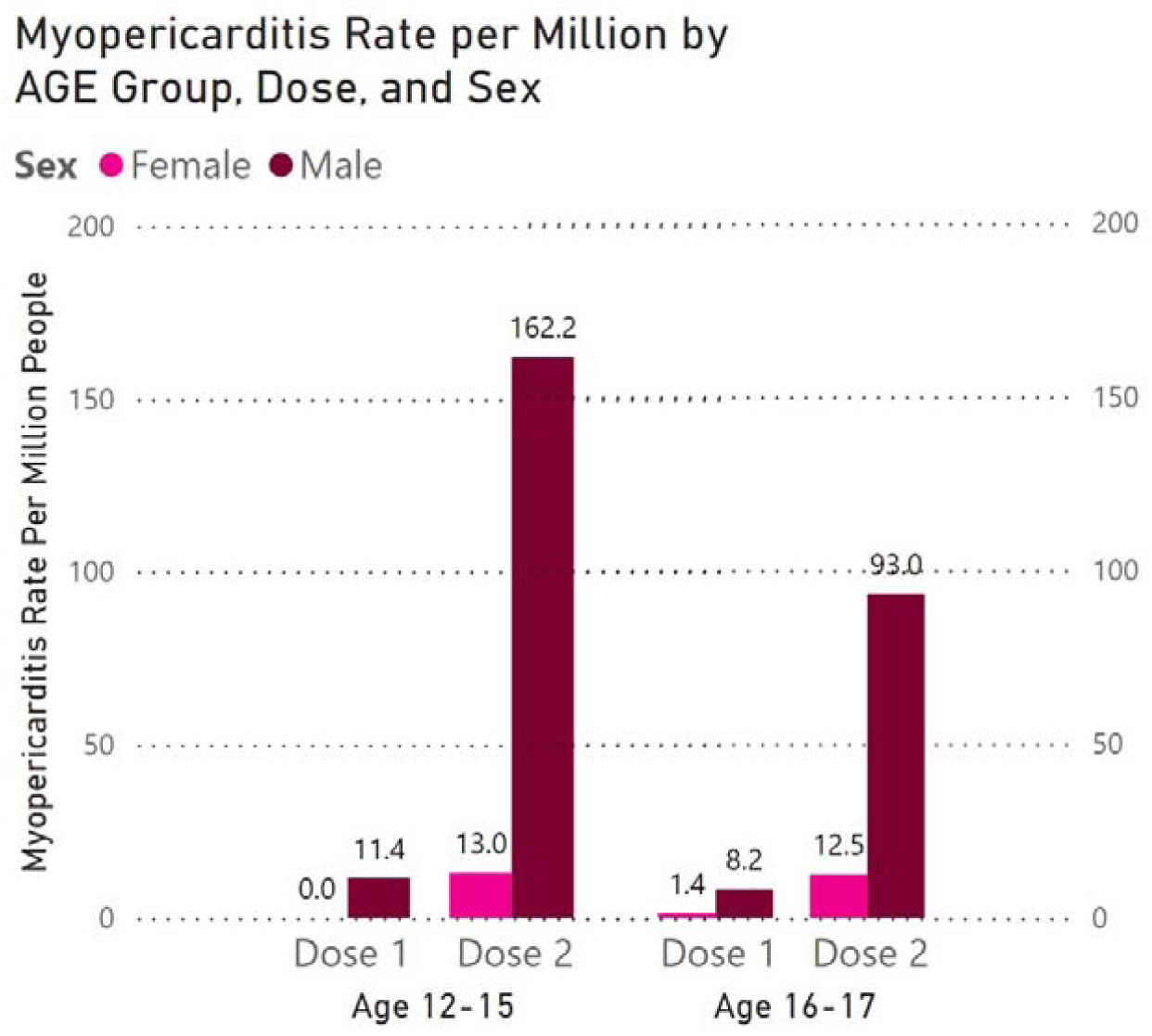
Cardiac Adverse Event (CAE) rate per million vaccinated persons, by age and sex and vaccination dose

**Table 1.** Cardiac Adverse Event (CAE) rates per million adolescents following vaccination doses 1 and 2, by age and sex.

|  | **Females (n=23)** |  | **Males (n=230)** |  |
| --- | --- | --- | --- | --- |
|  | Dose 1<br>(95% CI)<sup>a</sup> | Dose 2<br>(95% CI)<sup>b</sup> | Dose 1<br>(95% CI)<sup>a</sup> | Dose 2<br>(95% CI)<sup>b</sup> |
| **12-15 years** |  |  |  |  |
| CAE Criteria met | 0 | 8 | 21 | 100 |
| Denominator* | 1,834,687 | 616,511 | 1,834,687 | 616,511 |
| CAE Rate per million | **0**<br>**(0-0.20)** | **13.0**<br>**(5.6-25.6)** | **11.4**<br>**(7.09-17.5)** | **162.2**<br>**(132.0-197.3)** |
| **16-17 years** |  |  |  |  |
| CAE Criteria met | 2 | 13 | 12 | 97 |
| Denominator* | 1,471,878 | 1,042,863 | 1,471,878 | 1,042,863 |
| CAE Rate per million | **1.4**<br>**(0.17-4.9)** | **12.5**<br>**(6.64-21.3)** | **8.2**<br>**(4.21-14.2)** | **93.0**<br>**(75.4-113.5)** |

<sup>a</sup> Dose 1 denominator: at least 1 dose by June 11, 2021.
<sup>b</sup> Dose 2 denominator: fully vaccinated by June 11, 2021. Sex-specific denominator estimated by dividing total by 2.

Rates per million following dose 1 among males were: 11.4 per million (ages 12-15) and 8.2 per million (ages 16-17). Rates per million following dose 1 among females were 0 (ages 12-15) and 1.4 (ages 16-17). With respect to dose 1 and compared to CDC findings, 12–15-year-old boys were 1.4 times higher than the CDC, while girls this age had zero CAEs in our analysis. Among adolescents ages 16-17, our rate was 1.6 times higher for boys compared to the CDC’s rate [7] and the CDC’s reported rate for girls 16-17 post-dose one was 0.0 per million. [7]

The CAE cases in our investigation occurred a median of 2 days following vaccination, and 91.9% occurred within 5 days. The hospitalization rate in our report was 220/253 (86.9%) overall, with 111/129 (86.0%) in the 12–15-year-old cohort and 109/124 (87.9%) in the 16–17-year-old cohort. (Figure 2).

**Figure 2.**
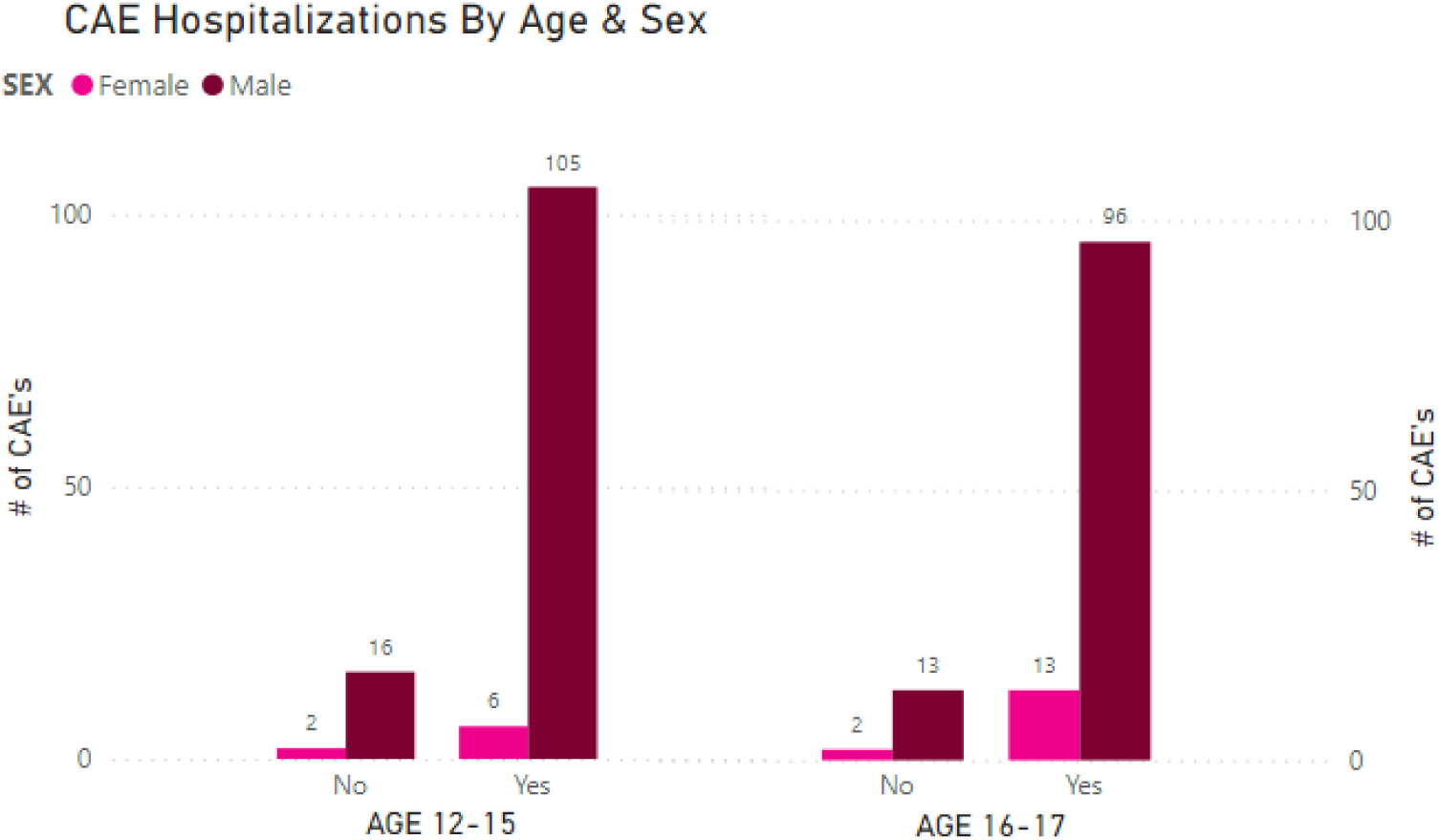
Total number of Cardiac Adverse Event (CAE) hospitalizations by age and sex

The median peak troponin T/I (Figure 3) (normal <0.1 ng/mL) was 4.5 ng/mL in boys ages 12-15 and 9.9 ng/mL in boys ages 16-17; for girls, the medians were 0.8 ng/mL and 7.0 ng/mL, respectively. Peak troponin values exceeded 2 ng/mL for 71% of cases age 12-15 and 82% of cases age 16-17. Figure 4 shows the time course of troponin increases after vaccination: 183/208 (86.7%) measured as elevated within 4 days. For the 216 cases for which the dose number was available, 30 (14%) occurred after dose 1 and 186 (86%) occurred after dose 2. The 37 cases with an unknown dose number were assigned proportionately to the overall distribution among the cases with a known dose number: 5 were assigned to dose one and 32 were assigned to dose two.

**Figure 3.**
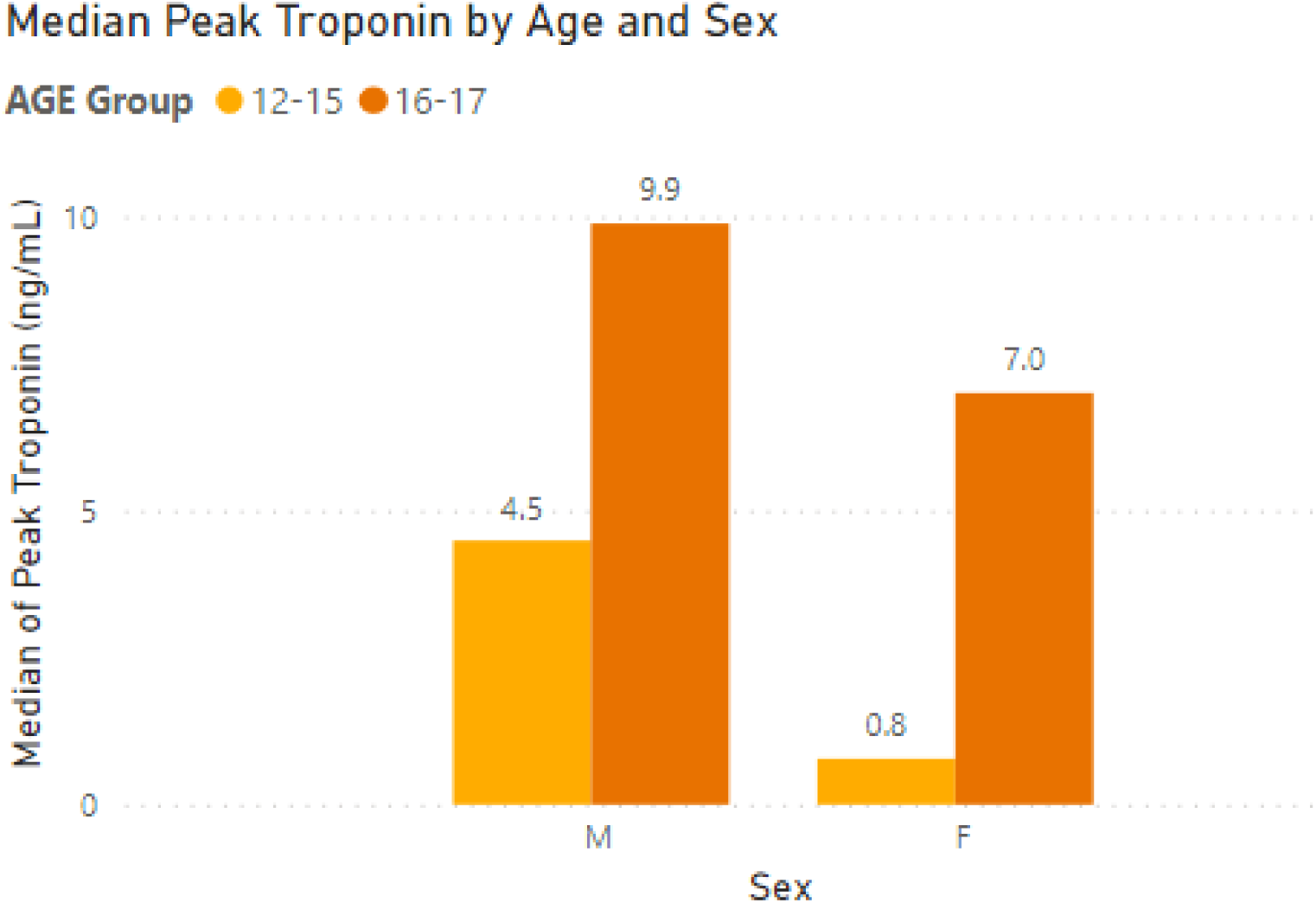
Median of Peak Troponin by age and sex in ng/mL

**Figure 4.**
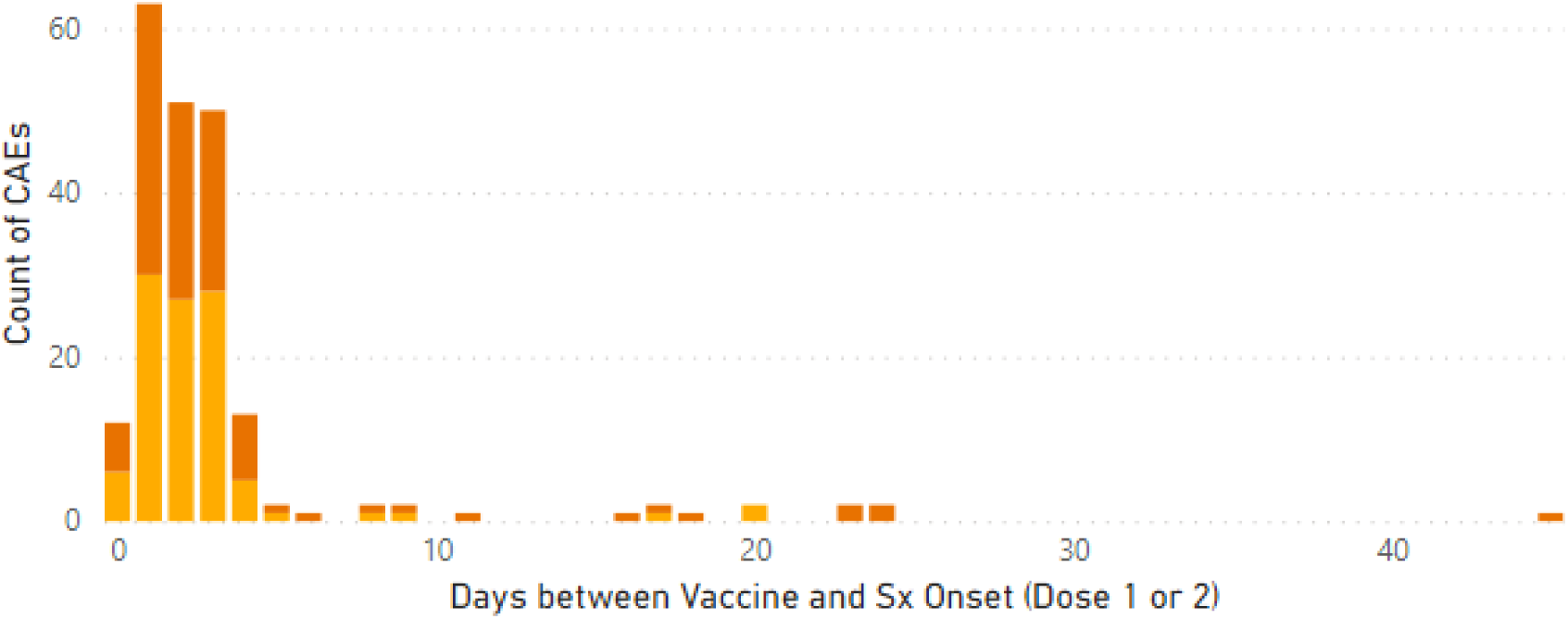
Symptom onset interval of Cardiac Adverse Events in days following vaccination among recipients with elevated troponin, by age

**Figure 5.**
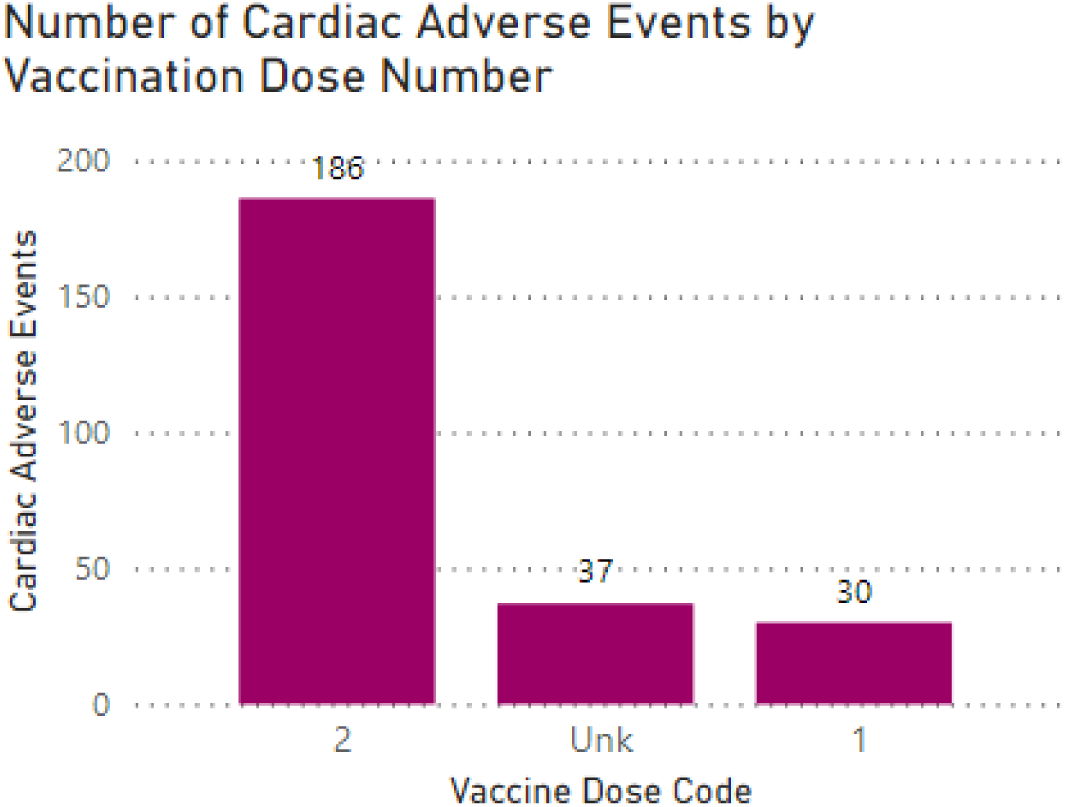
Vaccine recipients with cardiac adverse events by vaccination dose UNK= unknown

### COVID-19 hospitalization vs. vaccination harms

In the seven-month period of January 2021-July 2021, the rate of COVID-19 hospitalization among adolescents (ages 12-17) has ranged from a low of ≤ 4 per million weekly (July 2021) to moderate level (15 per million per week in mid-August 2021) and high (21 per million per week in January 2021) (Figure 6).[8,9] A healthy adolescent might expect a COVID-19 hospitalization risk of 44.4 per million over the next 120 days, assuming disease-related hospitalization prevalence stays at moderate levels (Figure 6, Figure 7). A child with at least one comorbidity might expect a disease-associated hospitalization rate of 210.5 per million per 120 days during times of moderate hospitalization, with a peak hospitalization risk of 294.7 per million per 120-days if rates surge to the same high levels as during the alpha wave (January, 2021).

**Figure 6.**
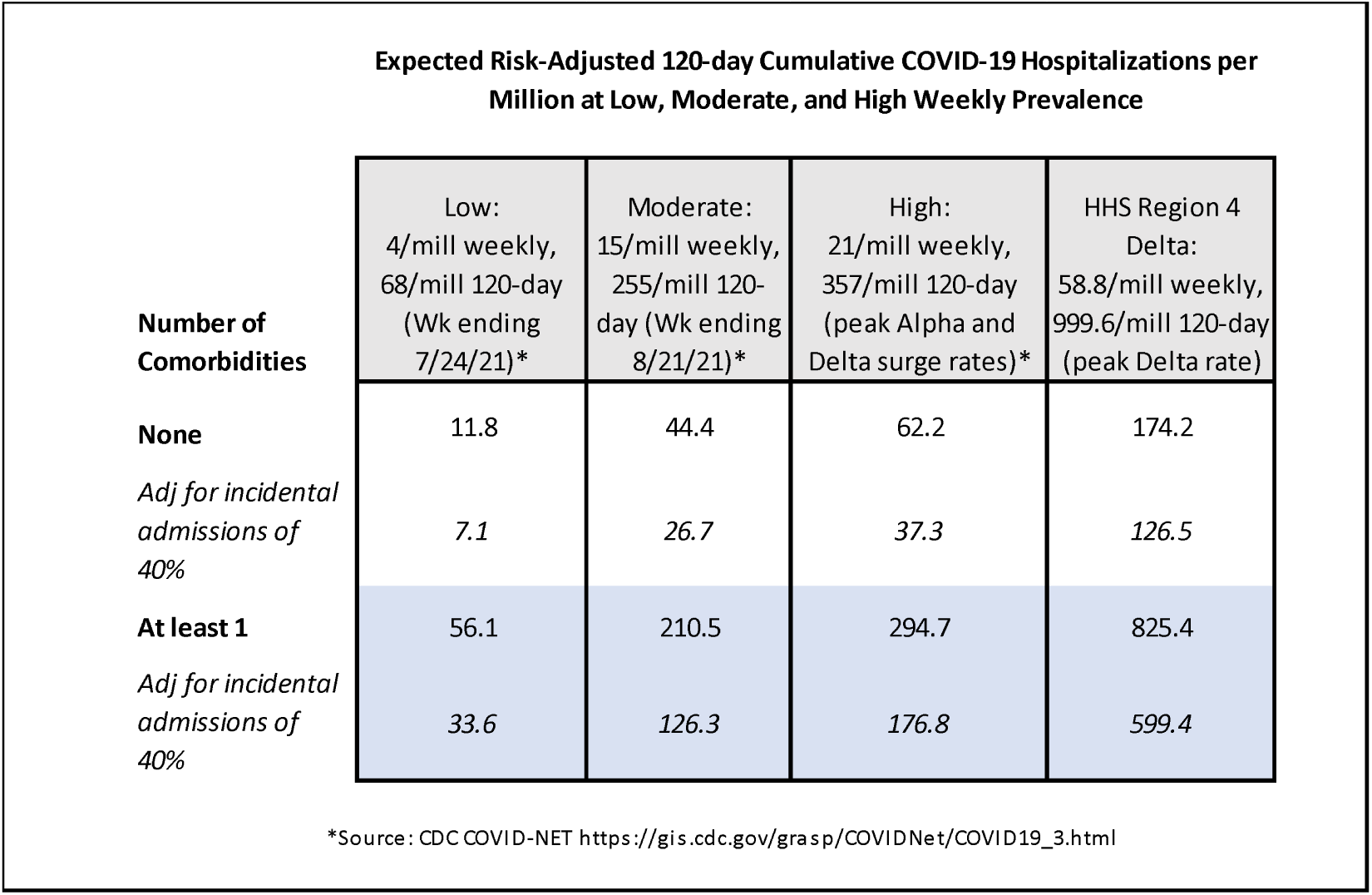
Harm-benefit analysis for second dose of mRNA vaccine vs. COVID-19 hospitalization for boys ages 12-17 in the context of disease incidence and presence of ≥1 comorbidity. *Darker shading denotes vaccine-associated risk of myocarditis equivalent to or exceeds disease-associated hospitalizations*.

**Figure 7.**
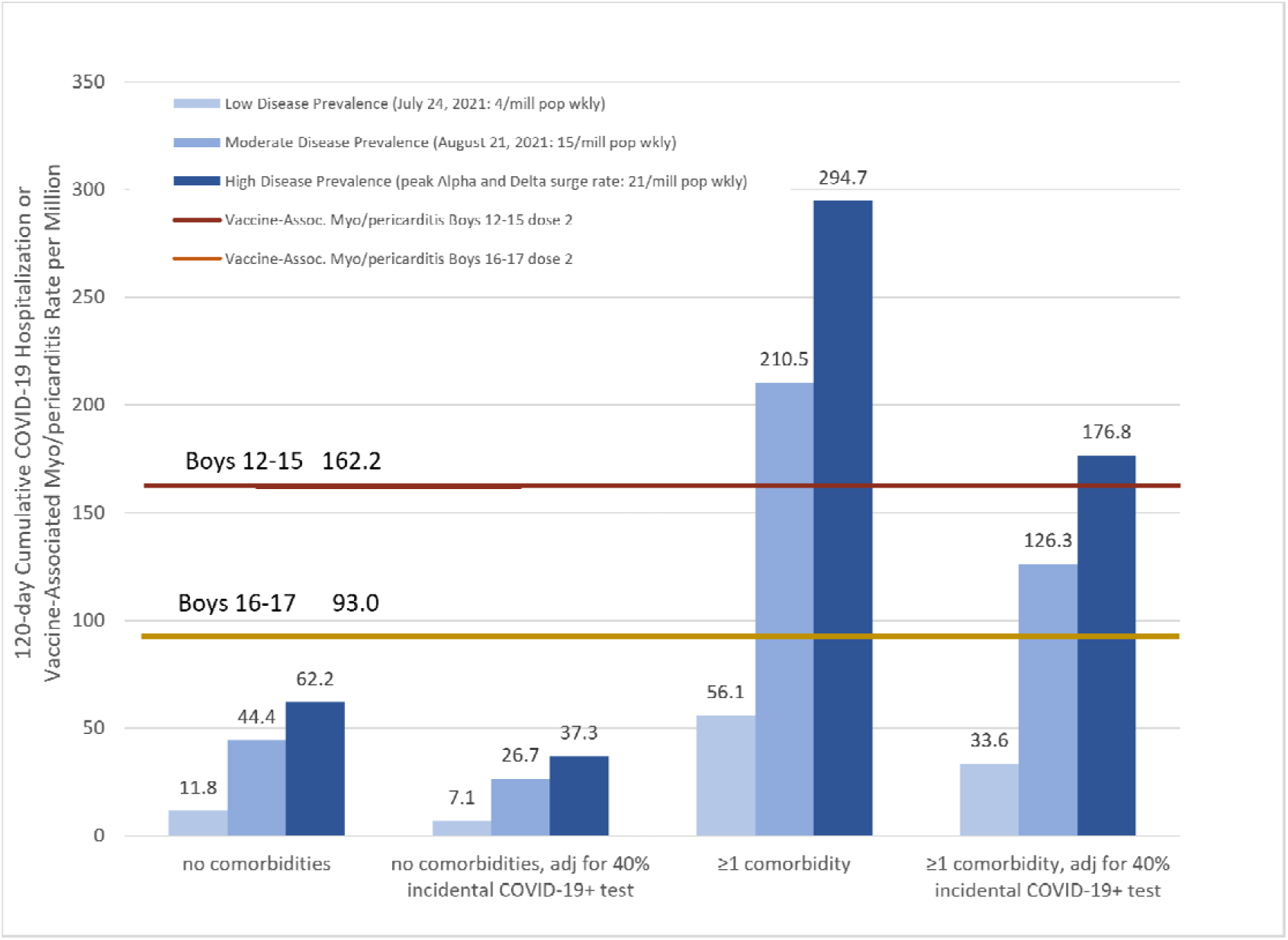
Harm-benefit analysis for second dose of mRNA vaccine cardiac adverse event (CAE) in boys by age vs. COVID-19 hospitalization risk in the context of disease prevalence and presence of ≥1 comorbidity.

At times of low adolescent COVID-19 hospitalization rates, such as in June 2021, a CAE from the second dose of an mRNA vaccine in a healthy 12–15-year-old boy was 13.7-fold more likely, at 162.2 per million, than the 120-day risk of COVID-19 hospitalization. The rate of post-vaccination CAE for boys 12-15 years without comorbidities (162.2/million) receiving their second vaccination dose exceeds their current 120-day COVID-19 hospitalization risk (44.4/million) by 3.7 times. In 16–17-year-old boys without comorbidities, the risk of post-dose two vaccination CAE exceeds their current 120-day hospitalization risk by 2.1 times (93.0/million vs. 44.4/million per 120-days). Our estimated risk of post-second vaccination dose CAE hospitalization for boys 12-15 without medical comorbidities (141/million; based on 86.9% hospitalization rate) also exceeds that of their 120-day COVID-19 hospitalization risk even at high hospitalization rates (Figures 6 and 7). For 12–17-year-old-boys with at least one medical comorbidity (Figure 6 and 7) their risk of post-vaccination CAE only exceeds their 120-day COVID-19 hospitalization risk at times of low hospitalization rates.

We also performed an adjusted analysis based on 40% of COVID-19 hospitalizations being due to another cause with incidental positive COVID-19 test during the hospitalization [9,13,14]. At current moderate incidence, the 120-day hospitalization risk for COVID-19 for a healthy child may be as low as 26.7 per million (Figure 6 in italics), twice the risk of a CAE after vaccine dose 1 in boys without comorbidities and after dose 2 in girls without comorbidities (Table 1). The risk for boys aged 12-15 with no comorbidities of CAE after dose two would be 22.8 times higher than their COVID-19 hospitalization risk at the adjusted low (7.1/million), 6.1 times higher for moderate (26.7/million) and 4.3 times higher at high (37.3/million) 120-day hospitalization rates (Figures 6 and 7). The risk for boys 16-17 with no comorbidities of CAE after dose 2 would be and 13.1 times higher than their COVID-19 hospitalization risk at the adjusted low (7.1/million), 3.5 times higher at moderate (26.7/million) and 2.5 times higher at high (37.3/million) 120-day adjusted hospitalization rates (Figures 6 and 7). For 12–15-year-old boys with one or more comorbidity, the vaccine-associated CAE risk following dose two is less than the adjusted COVID-19 hospitalization risk at times of high COVID-19 incidence. For 16– 17-year-old boys with one or more comorbidity, the CAE rate was below that of the adjusted COVID-19 hospitalization risks at both current (moderate) and high disease prevalence (Figure 7; Table 1).

## DISCUSSION

### Principal findings

The main finding of this study was the cardiac adverse event (CAE) rates of 162.2/million and 93.0/million post-Pfizer-BioNTech BNT162b2 vaccination dose two for the 12-15- and 16–17-year-old boys, respectively. Approximately 86% of these resulted in hospitalization for both age groups. We included a case-finding method in VAERS which utilized the symptom “chest pain” to identify adolescents for review of troponins, EKG/ECG and ECHO findings. We otherwise maintained the specificity of our analysis by requiring the same objective findings of cardiac injury used by the CDC to identify probable cases (Supplement 1) and excluded cases without sufficient objective evidence of cardiac-specific injury.

Using these broader search and inclusion criteria, we found post-vaccination rates of CAEs among adolescents aged 12-17 that exceeded the rates previously reported by the CDC[7] by 3.8 times in boys aged 12-15 and by 30% in 16-17-year-old boys. Our results show the risk of myocarditis depends heavily on sex and age, as it appears that young boys 12-15 have a greater than 12 times higher rate than girls. Due to a markedly increased risk for the youngest boys, prospective safety analyses would be most useful if stratified by ages 12-15 years and 16-17 years.

The highest troponin elevations were seen in the 16–17-year-old boys and girls. In the setting of cardiac symptoms, children with elevated troponin levels have a high likelihood of cardiac disease.[17, 18] Of note, the threshold for normal levels in children may be even lower than the 0.1 ng/mL used in adults.[17]

To contextualize the benefits of vaccination for adolescents, we chose, as the CDC did [2,3] to provide a benefit-harm analysis based on varying levels of COVID-19 hospitalization rates.[19] Our analysis not only considered circulating disease levels but also the presence of individual risk factors for severe disease. A weakness of our analysis was not being able to stratify hospitalization risks for children with and without natural immunity. The CDC estimated that on May 29th, 2021, that 36.2% [15] of all children had already been infected with SARS-CoV-2. This estimate was made 16 months into the pandemic, thus adjusting for the current time (19 months), the current estimate would be (19/16)*36.2% = 42.9% of children previously infected. Absolute infection-hospitalization rates for children with and without medical comorbidities and with and without history of infection have yet to be established.

In a recent report[20] of 15 patients aged 12-18 hospitalized with post-vaccination myocarditis, the clinical course was reported to be relatively benign, but 1/15 had abnormal echocardiogram on follow-up and 4/15 had ongoing symptoms post-discharge. The CDC reported [2] that 218 of the 323 (67.5%) cases of myo/pericarditis in vaccine recipients <30 years were known to have had resolution of symptoms. Another study[21] found 16/23 (70%) males with vaccination-associated myocarditis to have had resolution of symptoms within a week. Long-term sequalae of myocarditis is unknown; follow-up of this vaccine-associated condition is warranted.

COVID-19 has also been found to result in symptomatic myocarditis in 0.3% of collegiate athletes[22], but its rate in children post-COVID-19 infection has not been well described. The one existing study[23] is limited by an inappropriately small denominator due to apparent underestimate of COVID-19 disease incidence during the study period. The study[23] reported only six post-COVID myocarditis cases in boys ages 12-17 over the course of a year in a 60-million patient catchment area. Children have in general been spared from the worst effects of COVID-19. The reported mortality rate in England has so far been 2/million children [24,25], which may translate to around 6/million infections based on a prior infection rate of 1/3.[15] In the US, there have been 4404 multisystem-inflammatory syndrome in children (MIS-C) cases in a population of 74 million.[26] Long-duration of symptoms post-COVID-19 also appears less frequent in children than initially feared, with 1.8% of children with symptomatic COVID-19 experiencing at least one symptom two months after infection compared with 0.9% of negatively tested controls after one month.[27] A thorough understanding of vaccine safety is thus crucial in this age group, especially for children without known risk factors for severe COVID-19 and those with history of infection. This report only outlines rates associated with the Pfizer-BioNTech. A recent report from Canada [28] suggests a greater than 2-fold higher rate of post-vaccination myocarditis from the Moderna mRNA vaccine compared to the Pfizer-BioNTech mRNA vaccine. The most recent CDC update [7] also found Moderna to have higher rates of post-vaccination myocarditis than Pfizer-BioNTech in all male age groups except those 16-17 years and those 65 and older.

A concern about using VAERS for our data analysis is the risk of over-ascertainment of a safety signal due to the open access system. To address this concern, we aligned our inclusion criteria with the CDC’s case definition for probable myocarditis. Our rate of myocarditis post-vaccination of 93.0/million was more than two times lower than that reported by the FDA (200/million) [4] and yet 30% higher than the CDC. This suggests that both VAERS and CDC are providing an underestimate of the true incidence of this condition. Furthermore, the reports in VAERS reviewed for this study were of children with myocarditis *with* cardiac symptoms and not cases of incidental cardiac inflammation noted on imaging. It is thus unclear how large of an underestimate of CAE incidence this report provides.

We, like the CDC, used a 120-day COVID-19 hospitalization rate as a meaningful comparator to vaccination-related harms, but this type of harm-benefit analysis does not take into account any benefits the vaccine provides against transmission to others, long-term COVID-19 disease risk or protection from non-severe COVID-19 symptoms. With this in mind, the risk of CAE for a boy receiving his second dose of the vaccine is 2 to 6 times higher than the 120-day risk of hospitalization in boys 12-17 without underlying medical conditions. For boys with medical comorbidities, the 120-day COVID hospitalization rates are slightly higher than their rate of CAE if not adjusting for possible 40% overestimate of hospitalization rates [9,13,14], in which case the rate of CAE would be slightly higher than 120-day hospitalization [Figures 6 and 7]. As of the last week of August 2021, hospitalization rates for children ages 12-17 in the United States continued to approximate the moderate rate shown in Figures 6 and 7.[8] Current pediatric hospitalization rates are also a product of increasing natural infection and vaccination rates. Overall pediatric case hospitalization rates from COVID-19 have been estimated by the CDC [15] and American Academy of Pediatrics [19] to be 0.8-0.9%, irrespective of comorbidities and not adjusting for possible 40% overestimate of hospitalization rates [9,13,14]. This rate has remained consistent through August of 2021.

Given the nearly fivefold disparity in risk of hospitalization for adolescents with and without comorbidities, it is important that the benefits of vaccination be clearly weighed and conveyed in the context of the unique health risks of the individual and the household. The benefits of vaccination in previously-infected children should be further studied and a harm-benefit analysis performed. A history of SARS-CoV-2 infection may be found to provide similar or superior immunity to vaccination [29]. A recent study [30] found a 4-fold increased risk of post-vaccination myocarditis in those who had previously been infected with SARS-CoV-2.

In light of the vaccine-associated cardiac harms described in this report, further research as well as transparency about possible vaccine-related harms in relation to an individual child’s COVID-19 risks is indicated. Alternate vaccination types, dosing or strategies, such as those that take a history of infection into consideration, may eventually be found to be more appropriate in this age group.

## Conclusion

Our report found post-vaccination CAE rates following dose two of 162.2 and 93.0/million for boys 12-15 and 16-17, respectively. For boys with no underlying health conditions, the chance of either CAE, or hospitalization for CAE, after their second dose of mRNA vaccination are considerably higher than their 120-day risk of COVID-19 hospitalization, even at times of peak disease prevalence. The long-term consequences of this vaccine-associated cardiac inflammation are not yet fully defined and should be studied. In lieu of pediatric vaccination mandates, the US may: 1) consider gathering data on previous infection in this age group and/or 2) follow the example of Germany,[31] Sweden [32], Norway [33] and the WHO[34] and hold off on definitively recommending vaccination of low-risk children against COVID-19, or 3) offer one dose to adolescents as the UK has just announced [35] while more information about risks, benefits, harms and alternative dosing or vaccination strategies are studied and considered.

## Data Availability

Data used in this analysis are available at the link listed below.

https://bit.ly/CAEmRNA

---

**Supplement 1:**
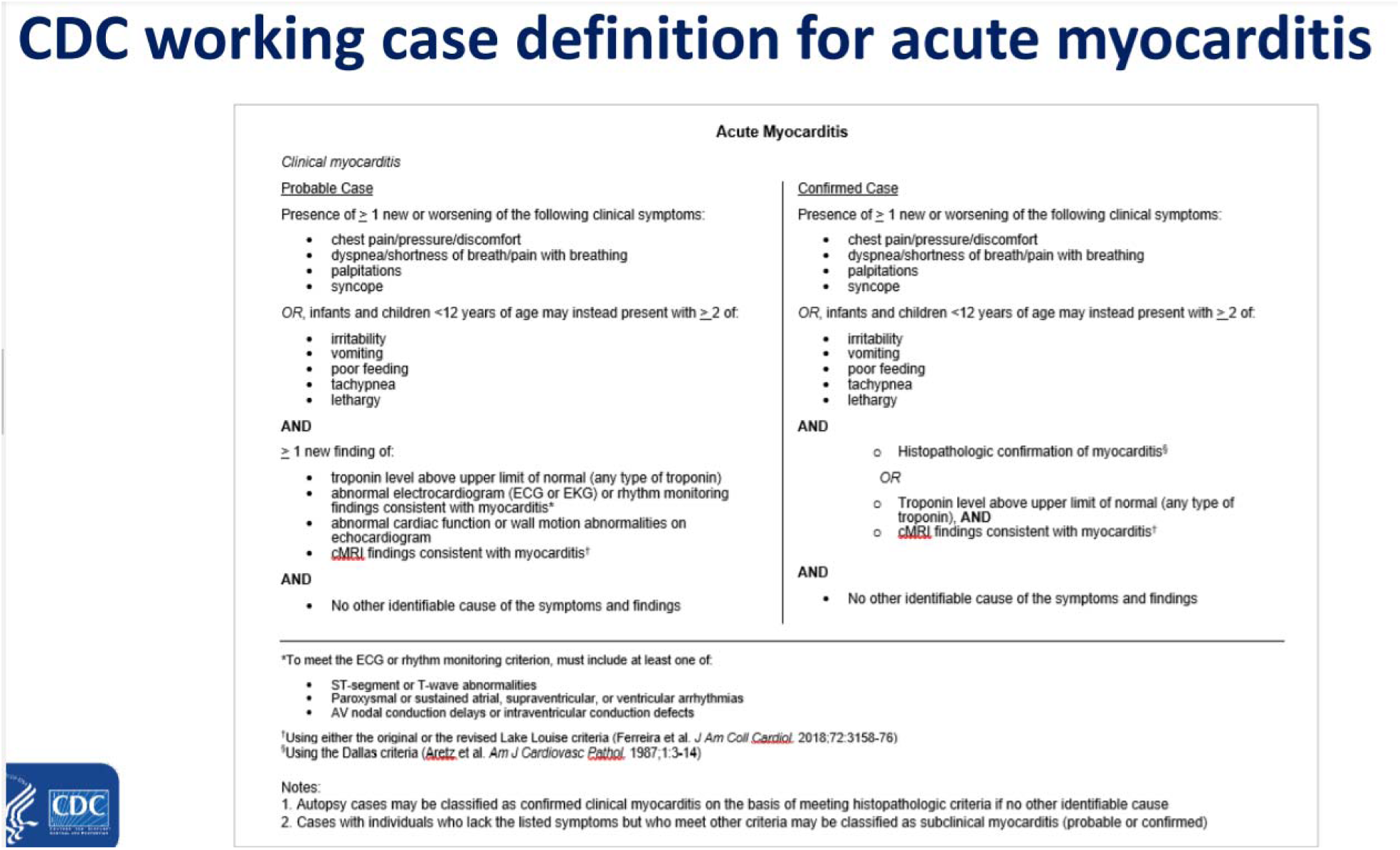
CDC working case definition for acute myocarditis

**Supplement 2:**
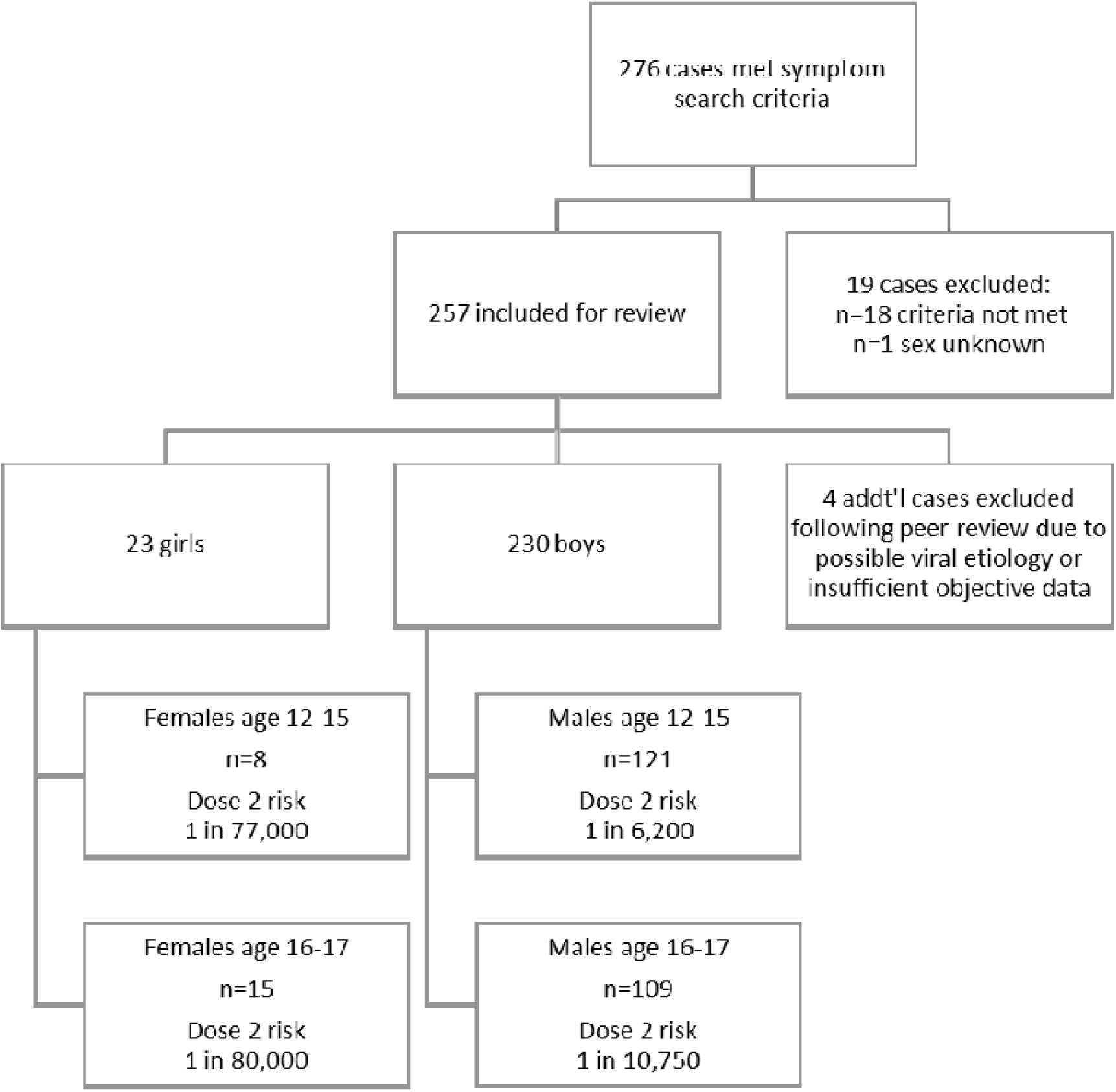
Study Inclusion Flowchart

**Supplement 3.**
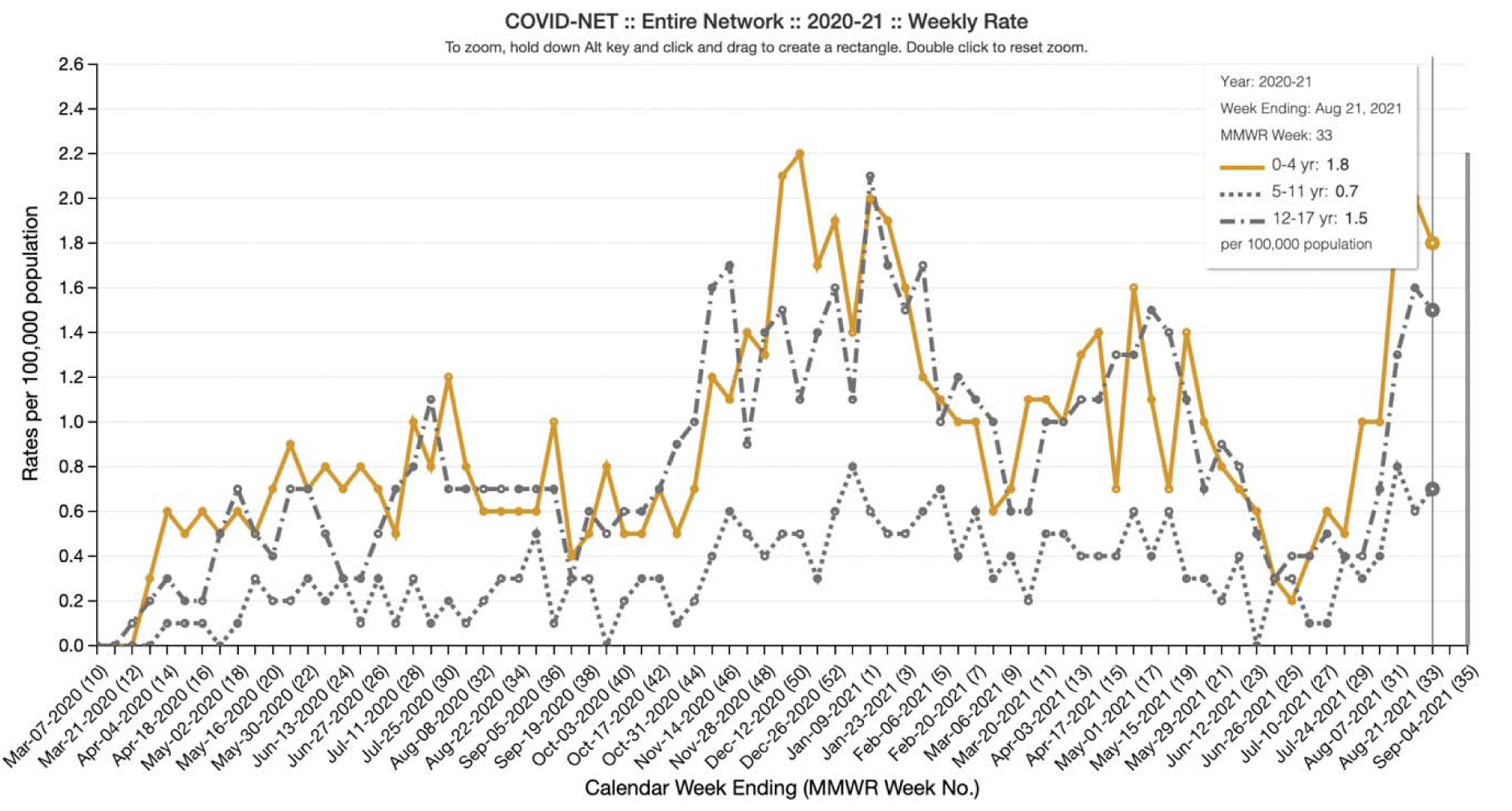
Pediatric COVID-19 hospitalization rates by age and week in the United States from COVID-NET [8].

